# Opioid Prescribing During Delivery Hospitalization Associated with Increased Postpartum Emergency Department Utilization

**DOI:** 10.1101/2020.10.26.20219923

**Authors:** Grace Bagwell Adams, Courtney R. Yarbrough, Samantha J. Harris, Amanda J. Abraham

## Abstract

**Background:** Empirical evidence shows postpartum women are prescribed opioids at high rates following both cesarean and vaginal delivery. Beyond this, little is known about opioid prescribing to postpartum mothers following delivery hospitalization and the resulting impacts for maternal health. Postpartum women may be particularly vulnerable to adverse health effects from inappropriate prescribing.

**Objectives:** This study examines the effects of postpartum opioid prescribing on emergency department (ED) utilization, controlling for delivery type and other maternal characteristics.

**Study Design:** We conduct a retrospective cohort study using 2008-2016 data from the Medical Expenditure Panel Survey (MEPS).

**Results:** We find 31% of women received a new opioid prescription following delivery, and that receipt of an opioid prescription following delivery hospitalization is associated with a 67% increase in the odds of an ED visit.

**Conclusion:** This study is one of the first to test the relationship between opioid prescribing and healthcare utilization among this population, contributing critical information to our understanding of implications for opioid prescribing after childbirth for postpartum women.

## Introduction

The opioid epidemic and maternal mortality are concurrent public health crises in the United States that result in premature death and decreases in life expectancy.(1-3) The intersection of these two crises is an area that merits the attention of policymakers, clinicians, and researchers. Adults of reproductive age are among those most affected by inappropriate opioid prescribing, opioid misuse, and opioid use disorder (OUD). Indeed, much attention has been garnered by documentation of the 400% increase in OUD among pregnant women between 1999 and 2014.(4-7) One target subpopulation that has been studied over the last few years with new focus is postpartum women who are opioid-naive that receive opioid prescriptions following delivery. Our study examines one of the lesser known pathways between these women receiving opioid scripts and their subsequent hospital utilization patterns.

In 2018, the American College of Obstetrics and Gynecology (ACOG) released the first ever guidelines for postoperative cesarean care, calling for a reduction in postoperative opioid prescribing. While ACOG recognizes the prevalence of opioid prescriptions for cesarean deliveries as a public health issue, it is important that we examine opioid prescribing for all births. Recent evidence shows that both cesarean *and* vaginal deliveries frequently result in receipt of an opioid prescription at delivery hospitalization.

Postpartum women who are opioid-naive may be particularly vulnerable to adverse health effects from opioids as they suffer from elevated rates of acute and chronic pain and are at risk of experiencing postpartum depression and anxiety. The potential link between opioid prescribing among opioid-naive women and their health care utilization and health outcomes is not well understood.

This paper examines the relationship between opioid prescribing and emergency department (ED) utilization after delivery hospitalization among postpartum, opioid-naive women. Using data from the Medical Expenditure Panel Survey (MEPS) from 2008 through 2016, we employ a logistic regression approach to estimate the association between receiving an opioid prescription following delivery hospitalization and ED use. Controlling for delivery type, insurance status, and a host of other demographic and socioeconomic characteristics, we find that receiving an opioid prescription is associated with a significant increase in the odds of ED use for postpartum women following delivery.

The literature on prescribing patterns for postpartum women has highlighted the prevalence of receiving an opioid prescription after birth and has shown important and significant differences in prescribing patterns based on delivery type--vaginal or cesarean delivery.(8) Regardless of delivery type, however, rates of opioid analgesic prescribing to postpartum women are high and have clinical and practical implications for both patients and their infant children. Research is needed to better understand the implications of opioid prescribing patterns in this population.

Understanding how prescription opioids are related to healthcare utilization and health outcomes can contribute new knowledge to efforts aimed at mitigating the opioid crisis *and* efforts to improve maternal health outcomes. Clinicians and policy makers, with greater understanding of the link between prescribing behavior related to both healthcare utilization and outcomes for postpartum women, will have increased ability to make more informed clinical decisions.

## Materials & Methods

To study the relationship between opioid prescribing and emergency department visits for postpartum mothers, we rely on publicly available data from the MEPS from 2008 through 2016. The MEPS is a nationally representative survey of non-institutionalized individuals’ health care utilization and spending conducted by the federal Agency for Healthcare Research and Quality. It follows survey participants during five data collection rounds over two-year, overlapping panels of approximately 16,000 individuals per panel.

We collect all childbirths from the Hospital Inpatient Stays files. Over the nine years of our study period, there were 3,569 live births. For those delivering mothers, we obtain socioeconomic, demographic, and insurance coverage variables from the Full-Year Consolidated Data files. We obtain data on mothers’ outpatient prescription fills from the Prescribed Medicines files and identify opioid prescriptions according to Multum drug category^1^. We create a dichotomous variable for a mother filling any opioid prescription in the birth round. To avoid confounding from opioid prescriptions resulting from an ED visit (unrelated to delivery), we omit opioid prescriptions linked in the data to non-delivery-related ED visits.

We limit our sample to mothers considered to be “opioid-naive” prior to delivery by excluding those who filled a prescription for any opioid in the survey round prior to delivery, in addition to births from mothers who received any medication treatment for opioid use disorder^2^ during the survey.(9) As such, we exclude births that occur during the first survey round in order to observe a full round of prenatal prescribing. We also exclude births that occur in the final (fifth) survey round in order to allow a sufficient length of time for post-delivery observation of our outcome variable. Finally, we exclude mothers who experienced a multiple birth (*i*.*e*., twins or triplets). Our final sample includes 2,245 births occurring in survey rounds two through four that met the other criteria described above.

For our outcome variables, we obtain records of ED visits for mothers from the Emergency Rooms Visits files, which contain Clinical Classification Codes (CCC) describing the reason for the ED visit. For each individual, we construct a dichotomous dependent variable indicating whether the mother had any ED visits during or after the month of birth. We exclude ED visits coded in MEPS as being prenatal- or delivery-related and those with CCCs related to pregnancy or delivery.^3^

We estimate descriptive statistics on the sample, disaggregated by those who received an opioid prescription and those who did not. Then, t-tests are estimated to examine the differences in means for variables of interest including receiving an opioid prescription, receipt by delivery type, and ED visits for those with and without an opioid prescription.

We then use a series of logistic regression models to measure the association between a postpartum mother filling an opioid prescription and the odds of a subsequent ED visit. Conditional on having an ED visit, we then estimate a model to predict the count of ED visits among postpartum women as associated with receipt of an opioid prescription. We control for a number of characteristics including whether the delivery was by cesarean section, mother’s age, race/ethnicity, educational attainment, health insurance status, poverty status, marital status, and family size.

We include fixed effects for year and geographic census region to control for secular time trends and for time-invariant regional characteristics. Because giving birth in an earlier survey round provides more opportunity for an ED visit to occur in the remainder of the survey period, we control for the number of months from delivery to the end of the survey. All analyses were conducted in Stata 15 using the MEPS-provided sample weights to make the results nationally representative. The study was based on de-identified data from a U.S. government public use dataset and was therefore exempt from IRB approval.

### Limitations

This study has several limitations of note. First, the small sample size constrains the predictive power in model estimation. Second, visiting the ED after delivery is a relatively rare event, thus the sample size is additionally constricted by the rarity of observing an ED visit. Third, we do not observe the timing of the ED visit--the observations are not at the patient-day level. Rather, we observe women in the birth month and in subsequent survey rounds after the birth month. We did, however, conduct sensitivity analyses to limit ED visits to the month after birth and then through the end of the survey participation period. Results are consistent across specification. Fourth, a we do not observe delivery-related injuries; we do, however, control for delivery type.

## Results

Exhibit 1 displays summary statistics of the final sample, which includes data on mothers from 2,245 births—69% vaginal deliveries and 31% cesarean. Mothers were prescribed a post-partum opioid in 30.0% of cases. Approximately 12% of new mothers visited the ED during the survey after their delivery; however, among women who received an opioid prescription, 16% had a subsequent visit to the ED compared with 10% of women who did not receive an opioid (p<0.01).

Approximately 40% of women who received an opioid at birth were white compared to 31% of women who did not receive an opioid at birth (p<.01). Conversely, 30% of women who received an opioid at birth were Hispanic compared to 40% of births with no opioid prescription (p<.01). Mother’s age, some college or an Associate’s degree, and being insured were all significantly higher in births with an opioid prescription. Having less than a high school education, falling below the Federal Poverty Level, and smaller household size were significantly lower in births with an opioid prescription.

Exhibit 2 shows a comparison of means by delivery type, opioid prescription filled, and ED visit. The first graph shows statistically significant differences in the percentage of women with vaginal versus cesarean deliveries (p<.01). The second shows the percentage of women in the total sample who fill an opioid prescription and then disaggregates the sample by delivery type. The percentage of women who received an opioid prescription at birth was significantly higher for women delivering via cesarean (p<.01). Finally, the third graph shows the percentage of women who experience an ED visit for the total sample and by delivery type. While there is a difference in the percentage of women with an ED visit by delivery type, it is not statistically significant (p<.30).

Exhibit 3 presents the difference in the percentage of postpartum women experiencing an ED visit by those who received an opioid prescription versus those that did not. Women who received an opioid prescription were significantly more likely to have an ED visit than those who did not (p<.01).

Logistic regression results showing the relationship estimated between opioid receipt and probability of having an ED visit are shown in Exhibit 4. Among women who filled an opioid prescription after delivery hospitalization, there is a 1.69 increase in the odds of having an ED visit (p<.05).

Delivery type was not significantly associated with odds of an ED visit for postpartum women. Maternal age was negatively associated with odds of an ED visit (p<.05). The odds of an ED visit were higher for women with some college or an Associate’s degree (1.86, p<.05), relative to women with no high school degree or a GED. Finally, the odds of an ED visit were lower for women living in the West, South, and Midwest relative to those in the Northeast (p<.05).

A second model was estimated to test whether there was an association between receipt of an opioid and the number of times a postpartum woman visited the ED. Conditional on having an ED visit, we found no association between the receipt of an opioid prescription and the number of times a postpartum woman visited the ED. Finally, as a sensitivity analysis, we examined the quantity of opioids prescribed (as measured in morphine milligram equivalents, or MMEs) and also found that the MMEs were not a significant predictor of having an ED visit.

## Discussion

We find that filling an opioid prescription after delivery hospitalization is associated with a 1.67, or 67%, increase in the odds of postpartum, opioid-naive women visiting the ED after hospital discharge. Additional analyses showed no statistically significant effect for MMEs on ED utilization; likewise, conditional on an ED visit, receipt of an opioid prescription does not increase the number of ED visits for postpartum women. Notably, we do not find a significant association between delivery type and ED utilization.

This study builds on recent descriptive work that finds significant variation in opioid prescribing patterns among postpartum women by delivery type.(8, 10, 11) These studies highlight the prevalence of receiving a postpartum prescription; specifically, Badreldin et al. found that almost a third (30.4%) of women who delivered vaginally and 87% of women who delivered via cesarean received an opioid analgesic, respectively.(10) Contributing to the reliance on prescribing of opioids following delivery is the high cesarean rate in the United States. In 2015, there were 1.3 million cesarean deliveries—32% of all births—making cesareans the most common inpatient surgical procedure in the U.S. [25]

In April of 2019, a study published in the American Journal of Obstetrics and Gynecology found that prescription of opioid analgesics after delivery was associated with persistent opioid use, with marginal differences by delivery type.(12) This study showed new evidence that while opioids are prescribed at higher rates for cesarean section deliveries, it could be the initial opioid prescription that is a greater predictor of persistent opioid use rather than the cesarean section itself.(12-14) Osmundson and colleagues estimated that postpartum opioid prescribing was linked to 21,576 new persistent opioid users annually.(12)

Outside of the research on the opioid crisis, the clinical standard of care for new mothers includes little interaction with healthcare clinicians after giving birth. Most women receive a six-week postpartum check and then are released from the care of their obstetrician until it is time for their annual wellness exam a year later. Clinical standards of care and the lack of interaction with providers after birth stand in stark juxtaposition to the standard for newborns and infants through the first year. Infants have many points of contact with pediatricians in the first year through a standardized well visit schedule.

Sparse follow-up care for postpartum women potentially increases the risk for adverse physical and mental health outcomes. Opioid prescriptions could exacerbate this vulnerability, as women receiving an opioid prescription could be more likely to experience postpartum anxiety and depression. Empirical evidence has shown that opioid use is associated with depression; in addition to work that shows associations between OUD, depression, and anxiety as comorbidities. Given that women are at risk for postpartum depression and postpartum anxiety after birth, additional research is needed to examine whether opioid prescribing is associated with increased incidence of postpartum depression, postpartum anxiety, and OUD.

There are a number of avenues through which opioid prescriptions could influence the propensity for a postpartum woman to visit the ED. These include but are not limited to increases in postpartum depression or anxiety, sedation leading to accidents and injuries, lack of mental clarity leading to suboptimal decision making, or other adverse reactions to the opioid medication itself. Clinicians and policy makers weighing these potential risks against the known benefits of opioids should consider work such as a 2017 study by Bateman and colleagues that finds extensive opioid prescribing to women following a cesarean delivery reported no pain improvements related to opioid prescribing. Another study finds that a small number of women receiving post-delivery opioid prescriptions transitioned to long-term, potentially problematic opioid use, with the likelihood of long-term use increasing with substance use disorder risk factors [28].

Significant attention on the opioid crisis and maternal and child health has focused on pregnant women or their infants once born (e.g. Neonatal Abstinence Syndrome, or NAS), not on opioid-naive women at delivery and in the postpartum period. Our study initiates a new line of work to link opioid prescribing after birth to healthcare utilization among postpartum, opioidnaive women. As previously stated, recent work that has focused on postpartum women has focused on descriptions of prescribing patterns by delivery type and potential associations between opioid prescribing and opioid misuse, abuse, and persistent opioid use.

Our study is important because it addresses a more common scenario than the transition to persistent opioid use or abuse; that is, it examines the experiences of new mothers who receive an opioid pain reliever prescription during the short but vulnerable period immediately after birth. Much of the focus of existing literature is on the effect of opioid use during pregnancy on labor, delivery, and birth outcomes. While understanding the impact of prenatal opioid use is essential, it is also important to examine the potential effects of post-delivery opioid use on maternal health. Clinical and population health researchers have only begun to research new mothers and interactions between opioid prescribing and postpartum health. We call attention to the need for more work on the effect of prescribing on healthcare utilization and health outcomes in addition to OUD. These include but are not limited to behavioral health issues such as postpartum depression and anxiety.

## Conclusion

National crises in both opioids and maternal and child health merit focus in research, policy, and practice. Over the last decade, the opioid crisis and maternal and infant health outcomes (*e*.*g*. mortality and other adverse birth outcomes) have grown in magnitude, receiving significant media and research attention. The research examining the link between these two national problems needs more attention and empirical research. In particular, we need a deeper understanding of the causal links between opioid prescribing and maternal and infant health.

Over the last several years, meaningful strides have been made in research on the relationship between opioid prescribing and myriad outcomes of interest. Postpartum-specific work has contributed important descriptive analyses of opioid prescribing, with particular focus on the rate of prescription opioids received by delivery type. The current study extends this line of work by looking at the association between postpartum opioid prescribing and ED utilization for women who have given birth, finding significant increases in the odds of ED use for these women despite the type of delivery they experienced.

More research is needed on the direct link between opioids and healthcare utilization and health outcomes for postpartum women and their infants. Concurrent foci of postpartum research must look at postpartum women with OUD *as well as* opioid-naive women who are prescribed opioids post-delivery. The policy implications of these findings include the importance of adhering to recent clinical guidelines and recommendations from the American College of Obstetricians and Gynecologists for opioid prescribing for postpartum women. The findings of this research may help inform health care practitioners about the implications of opioid prescribing after childbirth and may support the development of alternative pain management strategies for new mothers.

## Data Availability

All data are publicly available at the link provided.

https://www.meps.ahrq.gov/mepsweb/

---

**Figure 1.**
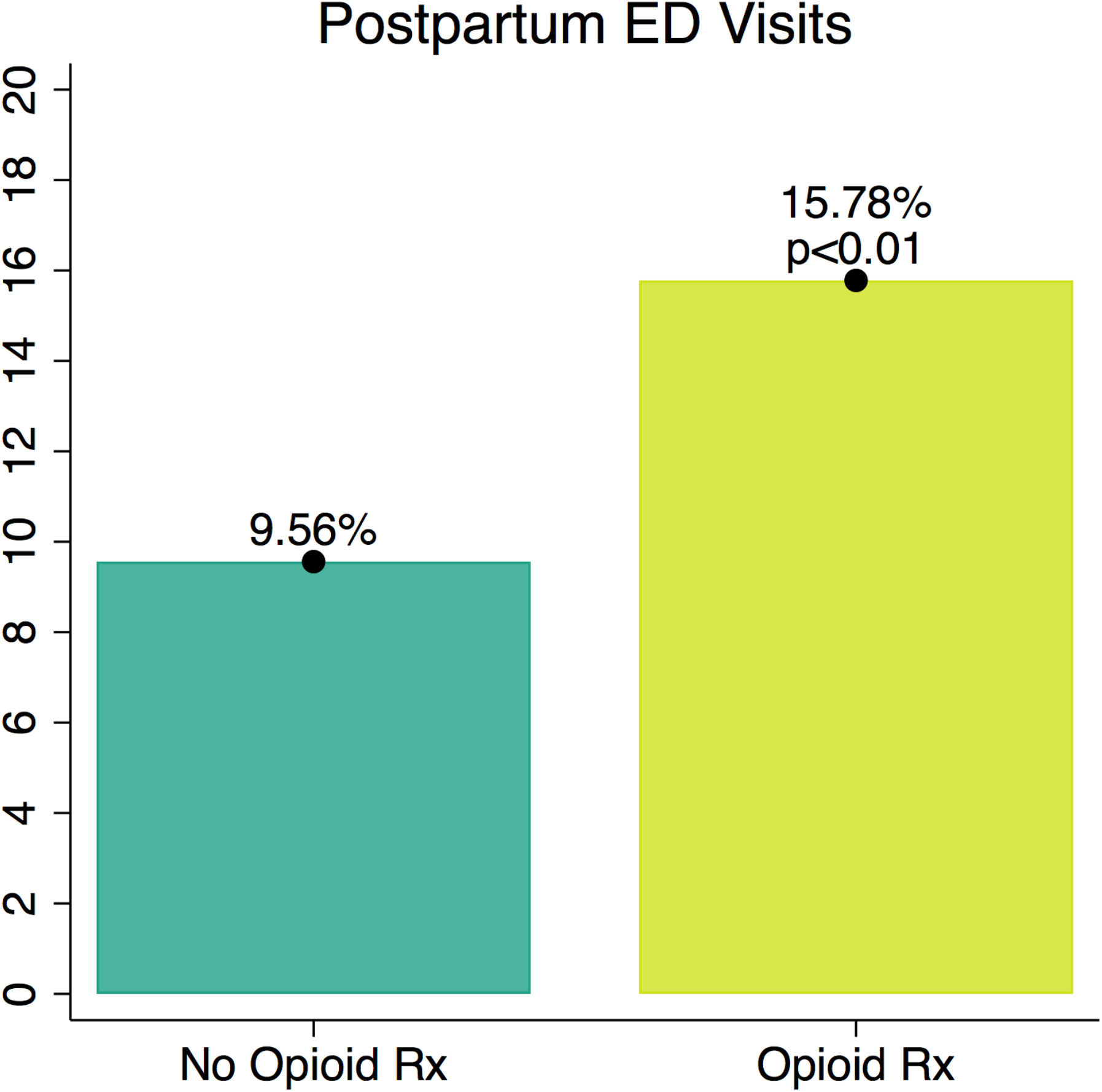

**Figure 2.**
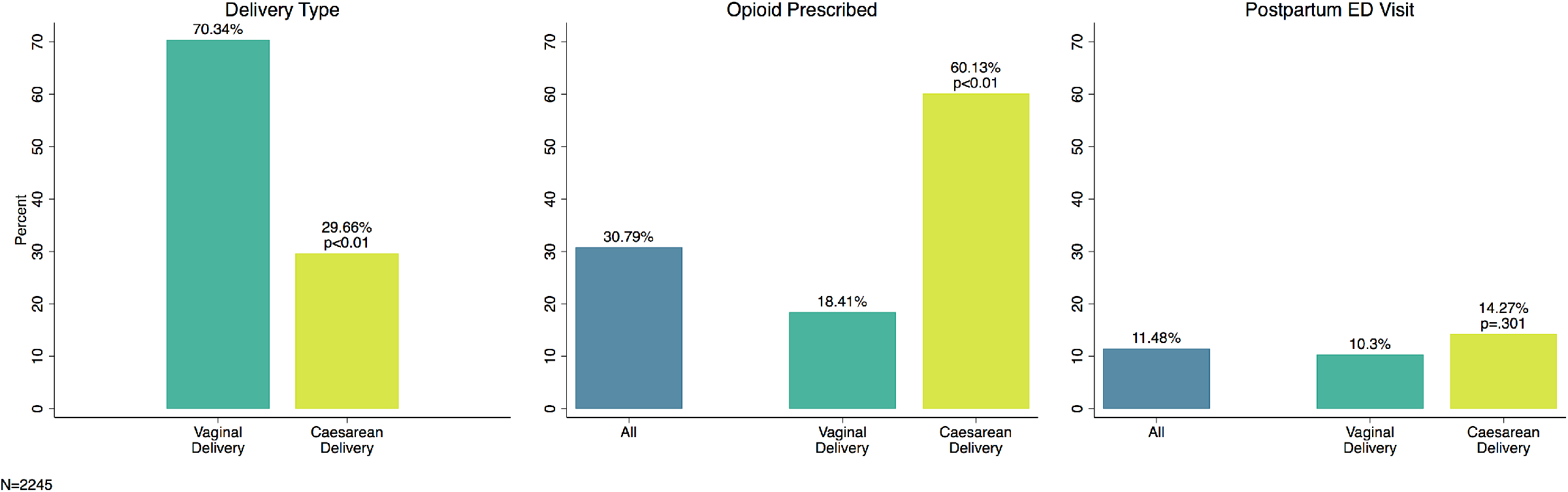

**Table 1.** Descriptive Statistics.

|  | All Births<br>mean (sd) | Births w/<br>Opioid Rx<br>mean (sd) | Births w/o<br>Opioid Rx<br>mean (sd) | T-Test by<br>Opioid Re-<br>ceipt |
| --- | --- | --- | --- | --- |
| **Dependent Variable:** |  |  |  |  |
| Any ED Visit | 0.12<br>(0.33) | 0.16 (0.37) | 0.10<br>(0.31) | ** |
| **Independent Variables:** |  |  |  |  |
| Any Opioid Rx | 0.30<br>(0.46) | 1.00 (0.00) | 0.00<br>(0.00) |  |
| Caesarean Delivery | 0.31<br>(0.46) | 0.58 (0.49) | 0.20<br>(0.40) | ** |
| White | 0.33<br>(0.47) | 0.40 (0.49) | 0.31<br>(0.46) | ** |
| Black | 0.20<br>(0.40) | 0.21 (0.41) | 0.20<br>(0.40) |  |
| Hispanic | 0.37<br>(0.48) | 0.30 (0.46) | 0.40<br>(0.49) | ** |
| Asian | 0.07<br>(0.26) | 0.07 (0.25) | 0.07<br>(0.26) |  |
| Other Race | 0.02<br>(0.15) | 0.03 (0.17) | 0.02<br>(0.14) |  |
| Mother's Age | 27.77<br>(6.06) | 28.15<br>(5.87) | 27.60<br>(6.13) | + |
| No HS Deg. | 0.22<br>(0.42) | 0.17 (0.38) | 0.24<br>(0.43) | ** |
| HS Deg./GED | 0.30<br>(0.46) | 0.33 (0.47) | 0.29<br>(0.46) | + |
| Some College/Associate's<br>Deg. | 0.24<br>(0.43) | 0.26 (0.44) | 0.23<br>(0.42) |  |
| Bachelor's Deg. or High-<br>er | 0.23<br>(0.42) | 0.24 (0.43) | 0.23<br>(0.42) |  |
| Insured | 0.82<br>(0.39) | 0.85 (0.36) | 0.80<br>(0.40) | * |
| Income <100% FPL | 0.40<br>(0.49) | 0.39 (0.49) | 0.40<br>(0.49) |  |
| Family Size | 4.38<br>(1.78) | 4.19 (1.66) | 4.47<br>(1.82) | ** |
| Married | 0.54<br>(0.50) | 0.55 (0.50) | 0.54<br>(0.50) |  |
| Mos. Birth-to-End Survey | 11.69<br>(5.13) | 11.65<br>(5.09) | 11.71<br>(5.15) |  |
| Northeast Census Region | 0.15<br>(0.36) | 0.12 (0.33) | 0.16<br>(0.37) | * |
| Midwest Census Region | 0.21<br>(0.41) | 0.23 (0.42) | 0.21<br>(0.41) |  |
| South Census Region | 0.36<br>(0.48) | 0.39 (0.49) | 0.35<br>(0.48) | + |
| West Census Region | 0.27<br>(0.45) | 0.26 (0.44) | 0.28<br>(0.45) |  |
| **Birth Years:** |  |  |  |  |
| 2008 | 0.14<br>(0.35) | 0.14 (0.35) | 0.14<br>(0.35) |  |
| 2009 | 0.12<br>(0.32) | 0.12 (0.33) | 0.12<br>(0.32) |  |
| 2010 | 0.11<br>(0.31) | 0.09 (0.29) | 0.12<br>(0.32) |  |
| 2011 | 0.11<br>(0.32) | 0.12 (0.32) | 0.11<br>(0.31) |  |
| 2012 | 0.12<br>(0.33) | 0.12 (0.32) | 0.12<br>(0.33) |  |
| 2013 | 0.12<br>(0.32) | 0.12 (0.33) | 0.11<br>(0.32) |  |
| 2014 | 0.10<br>(0.30) | 0.11 (0.31) | 0.10<br>(0.29) |  |
| 2015 | 0.09<br>(0.29) | 0.10 (0.30) | 0.09<br>(0.29) |  |
| 2016 | 0.09<br>(0.29) | 0.08 (0.27) | 0.09<br>(0.29) |  |
| Observations | 2245 | 674 | 1571 | 2245 |

*Source: Authors' analysis of births in the Agency for Healthcare Research and Quality's Medical Expenditure Panel Survey (2008-2016) among opioid-naïve women without multiple pregnancies.*
*Notes: ED = Emergency Department; Rx = Prescription; OR = Odds Ratio; HS = High School; Deg. = Degree; FPL = Federal Poverty Level; Mos. = Months*
Standard deviations in parentheses
Statistically significant differences in means by opioid receipt determined using the Student's T-test in Stata 15.
+ p<0.1, \* p<0.5, \*\* p<0.01

**Table 2.** Model Results.

|  | **Any ED Visit** |
| --- | --- |
|  | OR (p) |
| Any Opioid Rx | 1.69* (0.02) |
| Vaginal Delivery | *Ref. Cat.* |
| Caesarean Delivery | 1.17 (0.50) |
| White | *Ref. Cat.* |
| Black | 1.35 (0.25) |
| Hispanic | 0.63+ (0.08) |
| Asian | 0.43 (0.23) |
| Other Race | 1.15 (0.77) |
| Mother's Age | 0.96* (0.03) |
| No HS Deg./GED | *Ref. Cat.* |
| HS Deg./GED | 1.66+ (0.08) |
| Some College/Associate's Deg. | 1.86* (0.04) |
| Bachelor's Deg. or Higher | 1.11 (0.81) |
| Insured | 1.31 (0.33) |
| Income <100% FPL | 1.23 (0.33) |
| Family Size | 0.98 (0.76) |
| Married | 0.88 (0.64) |
| Northeast | *Ref. Cat.* |
| Midwest | 0.57* (0.04) |
| South | 0.62+ (0.07) |
| West | 0.48* (0.02) |
| Constant | 0.12** (0.00) |
| Mos. Birth-to-End Survey | Yes |
| Year F.E. | Yes |
| Survey Weights | Yes |
| Observations | 2,245 |
| F-Statistic | 5.14 |

Exponentiated coefficients
*Source: Authors' analysis of births recorded in the Medical Expenditure Panel Survey (MEPS), 2008-2016. All models conducted using the logit command in Stata 15 and the MEPS survey weights to make estimates nationally representa-*
tive.
Notes: *ED* = Emergency Department; *Rx* = Prescription; *OR* = Odds Ratio; *HS* = High School; *Deg.* = Degree; *FPL* = Federal Poverty Level; *Mos.* = Months; *F.E.* = Fixed Effects
p-values in parentheses
+p<0.1, \* p<0.05, \*\* p<0.01

## Footnotes

1 Narcotic Analgesics or Narcotic Analgesic Combinations

2 Methadone, buprenorphine, or naltrexone

3 Clinical Classification Codes 176-196

